# Identifying and managing problematic trials: a Research Integrity Assessment (RIA) tool for randomized controlled trials in evidence synthesis

**DOI:** 10.1101/2022.05.31.22275756

**Authors:** Stephanie Weibel, Maria Popp, Stefanie Reis, Nicole Skoetz, Paul Garner, Emma Sydenham

**Author notes:** **Correspondence to:** PD Dr. Stephanie Weibel, Department of Anaesthesiology, Intensive Care, Emergency and Pain Medicine, University Hospital Wuerzburg, Oberduerrbacher Str. 6, 97080 Wuerzburg, Germany.

## Abstract

Evidence synthesis findings depend on the assumption that the included studies follow good clinical practice and results are not fabricated or contain errors. Studies which are problematic due to scientific misconduct, poor research practice, or honest error may distort evidence synthesis findings. Authors of evidence synthesis need transparent mechanisms to identify and manage problematic studies to avoid misleading findings.

As evidence synthesis authors of the Cochrane COVID-19 review on ivermectin, we identified many problematic studies in terms of research integrity and regulatory compliance. Through iterative discussion, we developed a Research Integrity Assessment (RIA) tool for randomized controlled trials (RCTs). RIA was piloted and used in updating this Cochrane review. In this paper, we explain the rationale and application of the RIA tool.

RIA assesses six study criteria: study retraction, prospective trial registration, adequate ethics approval, plausible authorship, sufficient reporting of methods (e.g., randomization), and plausibility of study results. RIA was used in the Cochrane review as part of the eligibility check during screening of potentially eligible studies. Problematic studies were excluded and studies with open questions were held in awaiting classification until clarified. RIA decisions were made independently by two authors and reported transparently. Using the RIA tool resulted in the exclusion of >40% of studies in the first update of the review.

RIA is a complementary tool prior to assessing ‘Risk of Bias’ aiming to establish the integrity and authenticity of studies. RIA provides a platform for urgent development of a standard approach to identifying and managing problematic studies.

## 1. Introduction

Systematic reviews aim to identify all studies that meet the eligibility criteria for the review. Studies that challenge the principles of good clinical practice and scientific integrity can mislead and corrupt the findings of a systematic review, and hence mislead guidelines and official recommendations that use the reviews. Public health laws may be developed based on the findings of systematic reviews, which has been particularly important in responding to the COVID-19 pandemic.^1^ This causes a dilemma for review authors: Ioannidis et al. pointed out that authors should become aware that false and fatally flawed trials are very common in the field of medicine and suggests toning down the confidence in their conclusions.^2^ Avenell et al. found evidence that trials retracted due to misconduct distorted the evidence base including systematic reviews, meta-analyses, narrative reviews and clinical guidelines citing such trials and concluded that many of those guidelines, systematic or other reviews would likely change their findings if the affected trial reports were removed.^3^

The COVID-19 pandemic threw these dilemmas into sharp focus. We systematically synthesized the evidence for the Cochrane review “Ivermectin for preventing and treating COVID-19”.^4^ During the second pandemic year and after publication of the Cochrane review, several clinical trials were retracted due to critical concerns on trustworthiness of the studies.^5-8^ After their retraction, studies claiming to prove ivermectin’s huge beneficial effect for treating this disease remain considered in published evidence syntheses.^9-12^ Additionally, it has to be considered that even if evidence syntheses are retracted as well^13^ or corrected for such distortions^14,15^ their initial impact on future research, guidelines, clinical practice, public opinion and patient preferences is hardly to be reversed. Whilst most of the retracted studies actually did not meet our inclusion criteria, we felt uneasy about the full extent of the problematic study pool investigating ivermectin for COVID-19, and decided to develop a tool to help us identify studies that were potentially “problematic” in relation to whether they had been fabricated, data had been altered, or were not in accordance with good clinical practice.

We used the Cochrane definition of a ‘problematic study’ as “any published or unpublished study where there are serious questions about the trustworthiness of the data or findings, regardless of whether the study has been formally retracted. Scientific misconduct will not be the only reason that a study might be problematic; problems may result from poor research practices or honest errors.”.^16^ We drew on a few tools available (such as the ‘REAPPRAISED’ checklist for evaluation of publication integrity^17^ and the data extraction tool from the Cochrane Pregnancy and Childbirth Group^18^ that addresses various aspects of scientific integrity). Cochrane has published guidance to facilitate research integrity checks in the reviews it publishes,^19,20^ but these checks have not routinely formed part of evidence synthesis or guideline development processes to date.

In this paper we describe the Research Integrity Assessment (RIA) tool to assess randomized controlled trials (RCTs) and how we have used it. This provides a platform for urgent development and adoption of a standard approach to sifting out problematic studies. This is a moral duty so they are not included in systematic reviews used to inform guidelines or as information for health professionals or the public.

## 2. Methods

### 2.1 Development of the new research integrity assessment tool

We developed a tool for the assessment of research integrity of RCTs focusing on investigational medicinal products (IMPs^21-23^). The tool was developed prior to preparing the first update of the Cochrane review “Ivermectin for preventing and treating COVID-19” (Ref. Cochrane review update) and is guided by specific questions of how to identify and deal with problematic studies in the context of this systematic review. The tool was developed by a group of content experts who were either authors (SW, MP, SR, NS) on the Cochrane ivermectin review, part of the editorial process, or had previously developed strategies for dealing with problematic studies in Cochrane Review Groups (SW, NS, PG, ES). Critical and important study characteristics to assure research integrity of RCTs have already been discussed and considered in various other publications and places,^17-19^ and some characteristics are legally required, such as ethics committee approval.^24^ We used iterative discussions, piloting preliminary forms, and web-conferences among the expert group to agree on critical and important study characteristics and handling of identified problematic studies in this systematic review. The assessment tool was developed in an Excel-based format and contains questions to critical and important criteria that help to identify problematic RCTs when deciding on inclusion into a systematic review. The tool was not validated for other clinical questions and evidence syntheses.

## 3. Results

### 3.1 RIA: Critical and important study characteristics to assess research integrity of RCTs investigating IMPs

We achieved consensus on six domains considering critical and important study characteristics to assure research integrity of RCTs investigating IMPs based on adherence to good clinical practice and scientific integrity: Retraction notices, prospective trial registration, ethics committee approval and written informed consent, study authorship, sufficient reporting of methods regarding relevant eligibility criteria (e.g. study design/randomization), and plausibility of study results. In the following paragraphs, we introduce the Research Integrity Assessment (RIA) tool based on critical and important criteria of study characteristics summarized in key domains, explain their rationale as well as methodological guidance. Critical and important criteria of the RIA tool are summarized in Table 1. An Excel-based format of the tool with critical and important signalling questions to the domains, is available online (https://doi.org/10.5281/zenodo.6460339).

**Table 1.** Critical and important criteria for a Research Integrity Assessment (RIA) of RCTs investigating IMPs for evidence syntheses

|  | Domain 1: Retraction or expression of concern | Domain 2: Trial registration | Domain 3: Ethics approval | Domain 4: Study authorship | Domain 5: Methods | Domain 6: Results |
| --- | --- | --- | --- | --- | --- | --- |
| Critical criteria | Retraction of the study | Registry number not reported<br>Not prospectively registered | Ethics committee approval not available<br>Name and location of the ethics committee not reported<br>Ethics approval number not reported<br>Written informed consent not obtained | Inconsistency of authors' affiliations and countries the study is reported to have taken place in<br>Inconsistency in different parts of the article, e.g. country and location of study conduct | Insufficient reporting of the study design, e.g. randomization |  |
| Important criteria | Expression of concern published elsewhere | Inconsistency in details such as dates and study methods reported in the publication and in the registration documents | Ethics committee approval not obtained by a nationally recognised ethics committee as defined in the country's clinical trial regulations | Implausible number of authors for the study design (e.g. a single author article reporting a randomised control trial is unrealistic) | Baseline details reported not in sufficient detail to assess whether randomization worked properly<br>Incomplete or missing study flow diagram (optional) | Implausible number of patients with the condition recruited within the timeframe<br>Unrealistic response rate or loss of follow-up<br>Number of participants (e.g. women) in each group do not coincide with the reported randomisation method (e.g. block randomization)<br>Plagiarism<br>Excessive similarity or difference in the characteristics of the study participants between groups<br>Discrepancies between data reported in figures, tables, and text<br>Calculation errors<br>Implausible study results (e.g. massive risk reduction) |

#### Domain 1: Retracted studies or studies with published expression of concern

##### The problem

Journal editors should retract a published study if they have clear evidence that the findings are unreliable.^25^ Consequently, retracted studies should not be included in evidence syntheses as they can distort the evidence base.^3^

##### Assessment

Cochrane has published detailed guidance on how to search for retraction notices and handle retracted studies in Cochrane reviews.^19,20^ Retracted RCTs can be identified by review authors as such through a search for post-publication amendments in the systematic search for studies or on the <u>Retraction Watch Database</u> (http://retractiondatabase.org/RetractionSearch.aspx?). Retracted RCTs should simply be excluded. An expression of concern can be published by a journal to raise awareness of a possible problem in a published study ^25^ and may also announce a full retraction. For RCTs with an expression of concern, review authors should by default move these to the pool of RCTs awaiting classification until resolution of the pending concerns.^19^

#### Domain 2: Prospective trial registration

##### The problem

The WHO declares that registration of all intervention trials is a scientific, ethical and moral responsibility and expects that all clinical trials are prospectively registered in a WHO Registry Network approved registry.^26^ The WHO regards trial registration as the publication of an internationally-agreed set of information about the design, conduct and administration of clinical trials. These details should be published on a publicly accessible website managed by a registry conforming to WHO standards. Registries checking data as part of the registration process may lead to improvements in the quality of clinical trials by allowing identification of potential problems early in the research process.^26^ One of the minimum standards set out for trial registries in the International Standards for Clinical Trial Registries (Item 2.3) is that registries must obtain written third-party confirmation of a trial’s existence as part of the registration process.^27^ Prospective trial registration can be a proxy for trial quality as investigators and authors of high quality trials know and follow these responsibilities, and prospectively registered studies have been shown to be at lower risk of bias.^28^ The exclusion of non- and retrospectively registered RCTs in systematic reviews may shrink the study pool dramatically because there is poor compliance with trial registration^29,30^ despite the fact that it may be legally required as defined in clinical trial regulations world-wide. On the other hand, this approach could bring us closer to the truth about the effectiveness of an intervention and gives us more confidence in the conclusions of our systematic reviews.^31^

##### Assessment

Whether or not a RCT has been prospectively registered can be proven in the trials register. The trial registry number should be reported in the study publication.^32^ Prospective registration is defined as registration of a trial in a recognized national or international trials register before enrolment of the first participant.^27^ Due to the increase in trial initiation early in the COVID-19 pandemic, a delay between submission of a trial registration to and the actual publication on the register web site may have occurred. Extraordinary circumstances such as this pandemic, however, do not release investigators from submitting their registration prospectively or justify a total lack of registration. To avoid an unfair and unreasonable judgement, the registration’s first submission date should be considered and deemed prospective if occurring before enrolment of the first study participant. In case of doubt, review authors should contact the authors for the submission date of the trial protocol and the RCT should be moved to the ‘awaiting classification’ section. Review authors should exclude non-registered and retrospectively registered RCTs.

There is no empirical evidence that inclusion of only prospectively registered RCTs in a review guarantees the inclusion of trustworthy studies only,^19^ therefore, additional study criteria are critical and considered in the following.

Review authors should also search for any inconsistencies in details of study methods (e.g. study design, allocation, masking) reported in the publication and in the registration documents. In case of inconsistency, review authors should contact the investigators for clarification.

#### Domain 3: Adequate ethics approval

##### The problem

Adherence to ethical principles in clinical studies is compulsory to protect the dignity, rights and welfare of research participants.^24^ As such, all clinical trials involving human beings have to be reviewed by an ethics committee to ensure that the appropriate ethical standards are being upheld.^24^ It is further good clinical practice that the study investigators obtain written informed consent from all participants before randomization.^33^ Details on ethics approval and written informed consent are included in the WHO trial registration guidelines.^27^

##### Assessment

Review authors should seek a published statement on whether approval from an ethics committee was granted and this statement should include the name of the ethics committee granting the approval, and an approval number.^19^ A statement on whether written informed consent was obtained, or a justification for its absence, should be included in the publication or be provided by the study author upon request. Lack of such statements in the article does not necessarily mean that a study did not have an ethics approval or did not obtain participant’s written informed consent. Therefore, if a study did not report the name of the ethics committee and/or the approval number and information on consent, review authors should send a request to the authors and the RCT should be moved to the pool of studies awaiting classification until clarified. If the authors cannot provide any component of the above, the RCT should be excluded. Moreover, it should be assured that a nationally recognised ethics committee as defined in the country’s clinical trial regulations gave the approval. The ethics committee can be searched for on the <u>WHO list of national ethics committees</u> (https://apps.who.int/ethics/nationalcommittees/nec.aspx) or searching the national responsible authority’s list of recognised ethics committees. Unfortunately, there is no international list of responsible authorities available and an individual web search might be necessary. An English language summary of the specific regulations for some countries can be found on the <u>NIH Clinical Trials Regulation website</u> (https://clinregs.niaid.nih.gov/).

#### Domain 4: Plausible study authorship

##### The problem

Authorship confers credit and has important academic, social, and financial implications, but authorship also implies responsibility and accountability for published work.^34^ When authorship is abused, accountability and responsibility can be questionable and the potential for manipulated analysis and conclusions may increase. The domain ‘plausible study authorship’ is a proxy for whether it is plausible that the study has actually taken place with a focus on authors’ details and the location where the study was conducted. There are certainly many more details regarding authorship that may be important to address, e.g. author contributorship, ghost authorship, and funding details. However, when applying our tool, other review authors are welcome to examine studies in more extensive detail in this domain. The REAPPRAISED list examines research governance, authorship, and research conduct, and details (unfortunately without further guidance) can be found there.^17^

##### Assessment

Review authors should focus on the authors’ details and the location where the study was conducted. Review authors should check whether an article is authored by individuals with affiliations different from the country where the study was conducted, though information about trial sponsorship, ethics committee approval, funding and regulatory oversight included in trial registry details can assist in evaluating such cases. Inconsistencies in the article regarding different countries specified in different parts of the article or as compared to the trial registry entry should flag a study as ‘potentially problematic’. Finally, the review authors should check whether the number of authors is plausible for the study design. In its most extreme form, one single author article reported an RCT, and may indicate a fabricated study, since it is impossible for one person to have managed such a complex study design alone.^19^ If there are concerns regarding a study’s authorship, a request should be sent to the authors and the RCT should be moved to the pool of studies awaiting classification until clarified. If the authors cannot justify any component or inconsistency of the above, the RCT should be excluded.

#### Domain 5: Sufficient reporting of methods (for example, randomization methods)

##### The problem

RCTs that report very sparsely on their methods regarding relevant review eligibility criteria (e.g. study design/randomization) immediately raise alarms, particularly 10 when study authors provide insufficient information to be able to make an assessment of the risk of bias using the Cochrane Risk of Bias (RoB) tools 1 or 2.0.^35^ With incomplete methods the RoB assessment frequently returns to ‘unclear’ or ‘some concerns’ which is actually misleading. Conversely, study authors who applied deficient study methods or introduced bias into their studies could avoid a poor risk of bias rating simply by underreporting.

##### Assessment

The most critical criteria for domain 5 is a sufficient reporting on study design methods (e.g. randomization) in the study report. Review authors should consider sufficient reporting of methods mainly as clear evidence that the study was truthfully randomized. The method used for the randomization must be described and the process must lead to a random allocation of the participants. The sole designation ‘randomized study’ is not sufficient. Baseline details must also be provided in sufficient detail to estimate whether the randomization worked. If it turns out that a trial declared as ‘randomized’ in the article was not truthfully randomized, the review authors should exclude the study. Considering details on the study flow diagram such as the number of participants being randomized, receiving the intervention, and being analysed, is optional for the RIA, as those aspects will be covered by the Cochrane RoB tool 1 and 2.0 (and missing information may be adjudicated for with a high risk of bias judgement). Therefore, with domain 5 it should be assured that only RCTs with sufficient reporting of their methods are to be included in the study pool of the review, and studies with insufficient details are held in awaiting classification until the authors provide further details upon request.

#### Domain 6: Plausible results

##### The problem

There are alarming numbers of false data or zombie trials published each year^2,36^ and this has major consequences for the entirety of the health research ecosystem and for naïve systematic reviewers who assume that the published studies are real. How can systematic reviewers identify and deal with false data or zombie trials when looking at published articles? In-depth checks of baseline details and individual participant data are time consuming and require specific statistical training – and both may be limited for most systematic reviewers. Therefore, we achieved consensus on a few criteria of reported results in RCTs, which we consider to be warnings requiring further checks for plausibility.^37^

##### Assessment

Review authors should assess for plausibility 1) the number of patients recruited within the timeframe with the condition; 2) the response rate or number of participants lost to follow-up; 3) the number of participants (e.g. women) in each group coinciding with the reported randomization method (e.g. block randomization); 4) excessive similarity or difference in the characteristics of the study participants between groups; and 5) results that could be implausible (e.g. massive risk reduction, unexpected outlier data, unusual frequency of a rare outcome). Furthermore, review authors should note any data error (e.g. number of participants or events that did not add up), calculation error, and discrepancies between data reported in figures, tables, and text. In addition, when multiple reports of the RCT are available, review authors should check for overlap in text and data between published articles by the same or different authors without explanation.

When working through those criteria, and inconsistencies or implausible data are identified, the review authors should send an information request to the study authors to allow for comments and clarification. Until resolution, the RCT should be held in awaiting classification. This domain should be handled with care as for the above mentioned complexity and should not lead to exclusion of a trial without clear evidence. We suggest to not exclude a RCT for this domain alone, especially if there are no other concerns raised when using this tool – it should apply *in dubio pro reo*.

### 3.2 When should the RIA be used during evidence synthesis?

It is important to assess RCTs that pass the PIC(O) (participants, intervention, comparator, and (outcomes)) eligibility screening as early as possible for research integrity. Exclusion of problematic RCTs cleans the whole study pool, not only the estimated effects in meta-analyses and conclusions thereof, but also qualitative analyses, summaries of baseline characteristics, and conclusions regarding evidence gaps. Therefore, early exclusion of problematic RCTs or movement to the awaiting classification category of inconclusive RCTs is preferred.

### 3.3 How should the RIA be used during evidence synthesis?

In the hierarchical workflow of RIA through domain 1 to 6, three decisions on a study’s eligibility are possible at any hierarchical step from 1 to 6: RCTs may be either included, excluded or moved to awaiting classification (Figure 1). The first three domains, e.g. retraction, prospective registration, and ethics approval, can frequently be answered with certainty and direct decisions without sending an author request (e.g. for retraction, retrospective registration, verbal informed consent). For domains 4 to 6, a definite decision on a study’s research integrity is much more difficult to find and correspondence with the study authors can be necessary. For performance of the RIA, it has to be kept in mind that RCTs should be included only if there is no obvious doubt regarding any of the critical criteria included in the tool. Retraction, lack of prospective registration, lack of adequate ethical approval with informed written consent, implausible study authorship, lack of truthful randomization, or implausible study results should lead to exclusion of a RCT. If there is any inconsistency, insufficient information or serious concerns, requests should be sent to the authors. Each study author must have the opportunity to respond and clarify the questions. When authors do not respond or questions remain unanswered, the RCT should be held in the awaiting classification category. Whenever it is concluded from the decision on one domain that a RCT has to be excluded, the following domains no longer apply, can be omitted and do not have to be answered. Systematic reviews in a living mode have to re-evaluate all included RCTs and RCTs held in awaiting classification for published retraction notices or expression of concern for each update.

**Figure 1.**
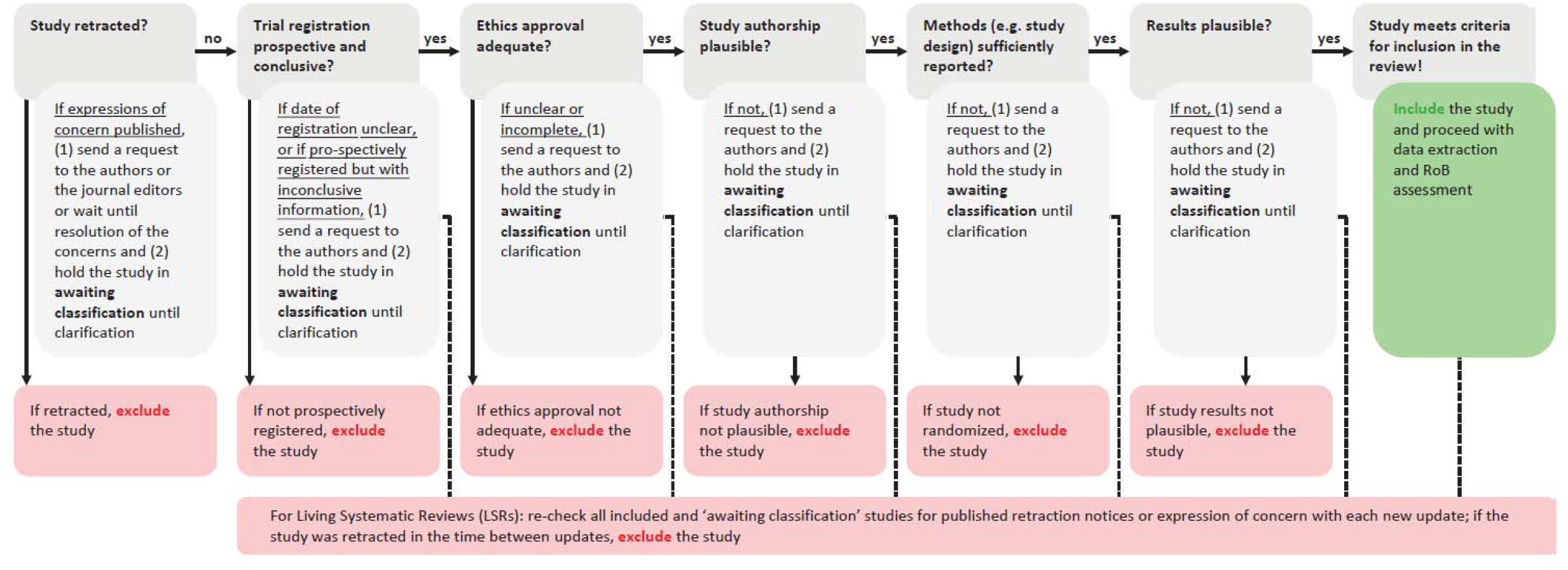
Hierarchical work flow and decision tree of the Research Integrity Assessment (RIA) tool. Potentially eligible RCTs identified during screening should be assessed for research integrity hierarchically considering domain 1 to 6. Retraction, lack of prospective registration, lack of adequate ethical approval with informed written consent, implausible study authorship, lack of truthful randomization, implausible study results should lead to exclusion of a RCT. Concerns with the RCT in any domain put the study in ‘awaiting classification’ and should lead to further investigations. If no concerns appear through all domains or could be clarified, e.g. in correspondence with study authors, the RCT meets criteria for inclusion in the review and can be processed further. In living systematic reviews, included RCTs and RCTs ‘awaiting classification’ must be reassessed for retraction notices.

The Excel version of the tool is based on critical and important signalling questions to the domains and includes columns to summarise a conclusion for each domain and an overall conclusion, which justifies the decision on research integrity and eligibility (https://doi.org/10.5281/zenodo.6460339). The tool offers a transparent way to document what the review authors have done (e.g. correspondence with the trial authors) and their judgements. For transparency, the table should be published as a supplement to the systematic review or deposited in an online repository with permanent digital object identifier (doi). The consequence of the RIA on the study pool should also be documented in the PRISMA flow diagram of the review using a new reason for exclusion ‘failed research integrity assessment’.

### 3.4 Working example: Cochrane ivermectin review

For the Cochrane ivermectin review, we evaluated 25 RCTs in our first review update (Ref. Cochrane review update): 14 included studies and three studies with results awaiting classification from the original review version^4^ were re-evaluated; and eight studies with results identified by the updated search were evaluated. In the review update, we excluded 11 out of 25 assessed studies, moved three to awaiting classification, and included eleven studies meeting all criteria for inclusion into the review update (Table 2, Supporting information 1).

**Table 2.** Research integrity assessment of RCTs applied to the Cochrane ivermectin review update. 25 RCTs were evaluated (14 included studies and three studies with results awaiting classification from the original review version, and eight studies with results identified by the updated search). The assessment took place hierarchically from domain 1 to 6 for all studies. Whenever it is concluded from the decision on one domain that a study has to be excluded, the following domains were not answered.

|  | Domain 1: Retraction or expression of concern | Domain 2: Trial registration | Domain 3: Ethics approval | Domain 4: Study authorship | Domain 5: Methods | Domain 6: Results |
| --- | --- | --- | --- | --- | --- | --- |
| Number of studies evaluated | 25 | 24 (-1) | 15 (-9) | 15 (-0) | 15 (-0) | 14 (-1) |
| Result | 1 retracted study (awaiting classification 1st review); identified via retraction watch database and post-publication amendment | 9 studies without prospective trial registration (6 included and 1 awaiting classification 1st review, 2 updated search); 4 non- and 5 retrospectively registered | 1 study posted results on a trials register without reporting ethics approval and number (awaiting classification 1st review); all studies with written informed consent | 2 studies posted results on a trials register and could not be assessed (1 awaiting classification 1st review, 1 updated search) | 1 non-randomized study (included 1st review); 3 study authors did not report/clarify sufficient detail on randomization (1 awaiting classification 1st review, 2 updated search) | 3 study authors did not clarify questions on study results (1 awaiting classification 1st review, 2 updated search) |
| Decision/Action | 1 study excluded<br>24 studies without concerns | 9 studies excluded<br>15 studies without concerns | no study excluded<br>1 study awaiting classification<br>14 studies without concerns | no study excluded<br>2 (+1) study awaiting classification<br>13 studies without concerns | 1 study excluded<br>3 (+1) study awaiting classification<br>11 studies without concerns | no study excluded<br>3 (+0) study awaiting classification<br>11 studies without concerns |

The most frequent reason for exclusion was a lack of prospective trial registration in nine studies; one study was retracted and another turned out not to be a randomized trial. The RIA excel sheet was published as a supplement to the updated Cochrane review and deposited in an online repository with permanent digital object identifier (doi, Ref.).

## 4. Discussion

As the number of systematic reviews being part of or being considered in evidence ecosystems continues to grow, we believe ensuring research integrity of studies included in the study pool of a systematic review is an approach that provides evidence closer to the unbiased truth and respects human rights. We hope the RIA tool, consisting of six domains to assess the research integrity of RCTs included in systematic reviews, will be viewed as a new transparent option to include the concept of research integrity in evidence synthesis and serves as a platform for urgent developments in this direction.

We believe the RIA tool represents a considerable expansion over previous efforts. The strength of our tool is that it was developed in accordance with recent Cochrane guidance on identifying problematic trials^16,19^ using an international team of (Cochrane) editors and content experts in systematic reviews methodology, clinical trial regulations, and evidence ecosystems. Using this tool for evaluation of research integrity of potentially eligible RCTs should be performed as part of the review‘s eligibility screening. Documentation of reviewers’ decisions on in- and exclusion will be transparent for evidence users increasing systematic reviewers’ accountability. We view that we need to implement an integrity assessment process quickly, but see this tool as the first version that can be adapted, modified, tested and validated over time.

This RIA tool is not a ‘Risk of Bias’ assessment. Critical aspects of research integrity assessed with the RIA are not considered in established RoB assessments such as the Cochrane RoB 1 or 2.0 tool. RIA aims to establish the integrity and authenticity of the studies. Cochrane’s research integrity department points out that the Cochrane RoB tool operates on the basis that the data are true.^38^ Research integrity of the trials is acknowledged as being a separate issue that should be handled prior to RoB assessment.

There may be several limitations to our approach. This tool was not validated with a larger sample of systematic reviews. The example presented here was used to develop the RIA tool performed on RCTs included in the ivermectin review for COVID-19. However, using this tool on trials in standard evidence synthesis work, where data are used from trials conducted in any country at any point in time, will not be as straightforward as in this example. Depending on the clinical question, in some cases, the majority of trials may have been conducted before trial registries even came into existence. Using this tool on trials is challenging in these circumstances, as there may not be any public records of trial governance, and alternative approaches may need to be developed, e.g. including older studies in secondary analysis only. A larger group of experts with different backgrounds and a Delphi panel process could have led to a different final set of critical domains. Future application of this tool may lead to refinement or adjustment of some domains, e.g. for different scenarios with intervention studies which are not IMPs. Identification and handling of false data or zombie trials by in-depth checks of baseline details and individual participant data requires specific statistical training – and both may be limited for most systematic reviewers. Statistical experts for IPD analysis and detection of fabricated data may not be well represented by our sample of experts. Therefore, the checking domain for plausibility of study results may still be under development and may change when new methods for identification of false and fabricated data become available. Research in this area is still in its infancy. Bordewijk et al. recently published a scoping review on methods to assess research misconduct in health research and concluded that tools to investigate are rudimentary and labour-intensive, and automatic tools and routine validation of these methods is needed.^37^ We plan to apply the RIA tool to a larger sample of systematic reviews to verify performance. With these findings, we plan to further develop the tool within the next three years in a Delphi process with a subsequent update.

We believe that this tool and the ivermectin review example has important teaching capabilities to raise awareness of the concept of research integrity for systematic reviewers and clinical study investigators. Clinical trial regulations are designed to ensure patient safety and must be adhered to. It is unfortunate that research was identified in the context of the ivermectin review that does not meet the required standard, and the only way forward is to improve access to Good Clinical Practice training for everyone involved in clinical trials, health research and evidence synthesis worldwide, with the aim of achieving full compliance with the regulations. We hope that research integrity criteria will lead to a rethinking so that the results of lawfully conducted studies are given prominence rather than treating all data equally in evidence synthesis. In addition, we hope that ensuring research integrity of studies included in the study pool of a systematic review is an approach that helps to provide evidence closer to the unbiased truth and improves respect of human rights in evidence synthesis.

## Data Availability

All data relevant to the study are included in the article, uploaded as supporting information, or available in the Cochrane ivermectin review with link to a public, open access repository.

## Acknowledgements

We thank the author team of the Cochrane ivermectin review: Selina Schießer, Renate Hausinger, Miriam Stegemann, Maria-Inti Metzendorf, Peter Kranke, Patrick Meybohm.

## Contribution of authors

The Research Integrity Assessment tool was developed prior to the preparation of the first update of the Cochrane review “Ivermectin for preventing and treating COVID-19” and addresses specific questions of how to identify and deal with problematic studies in the context of this systematic review. All authors of this article drafted the tool. SW, MP, and SR applied the tool to studies eligible for the Cochrane review. SW wrote the first version of the manuscript and all authors edited. SW is the guarantor of the article.

## Highlights

### What is already known

Including problematic studies in terms of research integrity in evidence syntheses can lead to misleading conclusions, and harm human health. Cochrane has published guidance to facilitate research integrity checks in the reviews it publishes. However, these checks have not routinely formed part of evidence synthesis or guideline development processes to date.

### What is new

We developed the RIA (‘Research Integrity Assessment’), a tool to assess the integrity of research and adherence to good clinical practice reported in RCTs of investigational drugs for an update of a Cochrane COVID-19 review. The assessment uses signalling questions to identify problematic RCTs and is used when studies are being considered for inclusion in an evidence synthesis. Problematic studies are excluded or held in an awaiting classification category.

### Potential impact for Research Synthesis Methods readers outside the authors’ field

Ensuring research integrity of studies included in evidence synthesis is an approach that helps to provide evidence closer to the unbiased truth and improves respect of human rights in evidence synthesis.

