## Supporting information 1 for "Identifying and managing problematic trials: a Research Integrity Assessment (RIA) tool for randomized controlled trials in evidence synthesis"

**Research integrity check of RCTs applied to the Cochrane ivermectin review update**

For the Cochrane ivermectin review, we evaluated 25 studies in our first review update (Ref. Cochrane review update): 14 included studies and three studies with results awaiting classification from the previous review version^1^ were re-evaluated, and eight studies with results identified by the updated search were evaluated.

Application of the RIA led to exclusion of 11 out of 25 assessed studies, three studies were moved to awaiting classification, and eleven studies meeting all criteria for inclusion were included into the review update. The most frequent reason for exclusion was a lack of prospective trial registration in nine studies; one study was retracted and another turned out not to be a randomized trial. The consequence of the RIA on the study pool were documented in the PRISMA flow diagram of the review using a new reason for exclusion ‘failed research integrity assessment’ (Ref. Cochrane review update).

**Domain 1: Retracted studies or studies with published expression of concern**

For the Cochrane ivermectin review update (Ref. Cochrane review update), we checked all studies in the update search for post-publication amendments and checked the names of the first and the corresponding authors of potentially eligible studies for retraction notices in the Retraction Watch Database (<http://retractiondatabase.org/RetractionSearch.aspx?>). One study from the study pool, which was classified as awaiting classification in the original Cochrane review due to insufficient and inconclusive information describing the randomization method, was retracted few months later.^1,2^ The responsible journal published a retraction amendment, which was identified by the updated search.^3^ The trial has also been listed in the Retraction Watch Database.^4^ A group of independent scientists showed that the individual patient data set of the study consisted of blocks of details from 11 patients that had been copied and pasted repeatedly – suggesting many of the trial’s apparent patients didn’t actually exist.^5^ Considering this issue in domain 1 of the presented tool, the retracted study was consequently excluded in the update of the Cochrane review (Ref. Cochrane review update).

**Domain 2: Prospective trial registration**

For the update of the Cochrane ivermectin review (Ref. Cochrane review update), nine of 24 non-retracted studies (37.5%) were excluded due to lack of prospective registration. Three included studies^6-8^ and one study awaiting classification^9^ from the previous review were excluded when reassessment revealed that they were not registered in any national or international study registry. Three included studies from the previous review^10-12^ and two studies with newly identified full-text publications^13,14^ were retrospectively registered; i.e. the date of first enrolment of participants was after the date of first protocol submission to the trial register. As described in the main paper, we used the date of submission instead of the date first posted to exclude a possible delay in the registration process at this point in the pandemic.

**Domain 3: Adequate ethics approval**

For our Cochrane review update (Ref. Cochrane review update), we checked the ethics approval of the 15 non-retracted and prospectively registered studies. Thirteen studies reported their ethics approval in the publication and the two unpublished studies reporting results in the trial register did not. Ten of the 15 studies reported the approval number in the publication or the study protocol, four provided them upon request and for one study the authors refrained from providing any data/information until journal publication of their results. The 14 studies providing necessary information were approved by a nationally recognized ethics committee. We independently confirmed the national recognition of the named ethics committee using public records, which took approximately one hour per study. We used Google Translate to assist in navigating our online search to the responsible authority information. It was easy to find information for the studies conducted in India, as the national authority maintains an open repository of national ethics committee registrations.^15^ Other countries may wish to follow India’s lead in making this information easily accessible. All studies obtained written informed consent, as reported in their publication or upon request by the review authors. Therefore, ethics approval was adequate for 14 studies. Information was missing for the one study that wants to share data once the study is published.^16^ This study was moved to the awaiting classification category.

**Domain 4: Plausible study authorship**

For the Cochrane review update (Ref. Cochrane review update), 12 of the 15 not retracted and prospectively registered studies were published and listed between seven and 29 authors. There were no concerns regarding authors’ affiliation and country of study conduct for the published studies. One author of an unpublished study sent all the information and details on the study to the review authors. Two studies posting their numerical results on the trials registry could not be assessed for this domain.

**Domain 5: Sufficient reporting of methods**

For the Cochrane review update (Ref. Cochrane review update), nine of the 15 not retracted and prospectively registered studies reported their respective method used for randomization in sufficient detail. Authors of six studies were contacted for further details on randomization. Three studies were categorized as awaiting classification because the study authors did not respond or provided insufficient explanation or they referred to full details planned to be published in a future publication.^16-18^ Three study authors responded, two clarified the randomization method and the other one turned out to be a non-randomized trial. The author’s description of the alleged randomisation method revealed that actually an alternate allocation was performed, and the study was therefore excluded.^19^ Eleven studies reported baseline characteristics of participants in sufficient detail. One study author provided additional baseline details upon request and three authors did not respond.

As a note, the six studies that were included in the original Cochrane ivermectin review that had to be excluded for the update as they were not prospectively registered also insufficiently reported on randomization, allocation concealment or used a predictable method.^6-8,10-12^ This supports the notion that prospective registration is a good proxy for trial quality.^20^

**Domain 6: Plausible results**

For the Cochrane review update (Ref. Cochrane review update), eight of the 14 non-retracted, prospectively registered, and randomized studies showed plausible results. For six studies, requests were sent to the authors, three of them satisfactorily responded, and the other three studies were categorized as awaiting classification because the study authors did not respond, provided insufficient explanation or they referred to full details planned to be published in a future publication.^16-18^ However, this decision was not simply based on this domain’s assessment but mainly on missing information on the randomisation method, which as a critical signalling question alone leads to this categorization.
